# End to end stroke triage using cerebrovascular morphology and machine learning

**DOI:** 10.1101/2023.02.27.23286357

**Authors:** Aditi Deshpande, Jordan Elliott, Bin Jiang, Pouya Tahsili-Fahadan, Chelsea Kidwell, Max Wintermark, Kaveh Laksari

## Abstract

**Background:** Rapid and accurate triage of acute ischemic stroke (AIS) is essential for early revascularization and improved patient outcomes. Response to acute reperfusion therapies varies significantly based on patient-specific cerebrovascular anatomy that governs cerebral blood flow. We present an end-to-end machine learning approach for automatic stroke triage.

**Methods:** Employing a validated convolutional neural network (CNN) segmentation model for image processing, we extract each patient’s cerebrovasculature and its morphological features from baseline non-invasive angiography scans. These features are used to detect occlusion’s presence and the site automatically, and for the first time, to estimate collateral circulation without manual intervention. We then use the extracted cerebrovascular features along with commonly used clinical and imaging parameters to predict the 90-day functional outcome for each patient.

**Results:** The CNN model achieved a segmentation accuracy of 94%. The automatic stroke detection algorithm had a sensitivity and specificity of 92% and 94%, respectively. The models for occlusion site detection and automatic collateral grading reached 96% and 87.2% accuracy, respectively. Incorporating the automatically extracted cerebrovascular features significantly improved the 90-day outcome prediction accuracy from 0.63 to 0.83.

**Conclusions:** The fast, automatic, and comprehensive model presented here can improve stroke diagnosis, aid collateral assessment, and enhance prognostication for treatment decisions, using cerebrovascular morphology.

## INTRODUCTION

Each year 800,000 people in the US suffering a stroke, leading to 140,000 deaths (1). Critically reduced regional cerebral blood flow during an acute ischemic stroke (AIS) initiates brain dysfunction and, if left untreated, brain tissue death at a high rate of 1.9 million neurons every minute (2). Fast and accurate AIS diagnosis is essential for timely treatment to limit irreversible brain damage (3), and improve functional outcomes (4).

AIS triage consists of baseline patient assessment followed by imaging to rule out hemorrhage, localize vessel occlusion and identify salvageable tissue-at-risk. However, the current clinical process relies on the immediate availability of vascular neurology and neuroradiology expertise, which varies significantly across institutions (4,5). In recent years, software solutions have been developed for the automatic detection of large vessel occlusion (LVO) and estimation of the ischemic core and at-risk tissue (4,5) using features such as the difference between left versus right hemispheric average vessel density (4,6,7). These approaches, however, are limited by low sensitivity (63-85%) or low specificity (50-70%), despite achieving high sensitivity (83-92%) due to pre-existing cerebrovascular changes such as intracranial atherosclerosis and tandem occlusions. Developing fast, automatic and accurate methods to extract and analyze complex cerebrovascular morphology can improve stroke protocols even in smaller facilities without access to local expertise (4,8,9).

Acute reperfusion therapies (ART), including intravenous thrombolysis (IVT), and increasingly, endovascular thrombectomy (EVT), are used for emergent recanalization of the occluded vessels (2,10). Accumulating evidence has established the effectiveness of EVT in improving the ischemic core to tissue-at-risk (penumbra) ratio and long-term functional outcomes (2). Hence, accurate and rapid patient selection is critical yet demanding and is typically performed using a combination of clinical and imaging parameters. The most common parameters include patient baseline functional status, symptoms onset time, stroke severity, baseline non-contrast computed tomography (CT) scan or magnetic resonance (MR) imaging of the brain, non-invasive vascular imaging modalities CT and Time-of-Flight MR angiography (CTA and ToF-MRA), as well as more advanced perfusion imaging in selected cased (11). However, not all AIS patients are eligible for ART and patient response to treatment relies on patient-specific cerebrovascular anatomy that governs cerebral blood flow during ischemia and reperfusion (12). Data-driven outcome prediction of ART can assist stroke systems of care in facilitating the triage and transfer of AIS patients and decision-making by physicians, patients, and their families. Therefore, automatic and individualized response prediction to treatment and outcomes is gaining popularity (13,14).

The collateral circulation is a network of secondary vessels developed over time to maintain cerebral perfusion and prolong tissue survival during ischemia (15). Accordingly, the optimal treatment window varies between individuals, and a better-developed collateral circulation provides the patient more time to receive acute reperfusion therapies (16). Therefore, a validated and rapid assessment of collateral circulation and potentially other complex cerebrovascular features can tremendously impact patient selection for treatment (17,18). The collateral index (CI) is a metric to quantify the extent of collateral circulation development. It has been shown to significantly impact patient recovery after recanalization and long-term functional outcomes (15). However, manual CI assessment is time- and labor-intensive and not incorporated into the routine stroke triage for patient selection and outcome prediction (19).

Machine learning (ML) algorithms have been increasingly used in recent years to improve multiple aspects of stroke care, including diagnosis, treatment, and outcome prediction (20). Many previous attempts have been limited by the variety and reliability of their input, which typically includes parameters similar to those used for EVT patient selection, such as the National Institute of Health Stroke Scale (NIHSS) (21) and the Alberta Stroke Program Early CT Score (ASPECTS) of the baseline non-contrast head CT scan (22). However, only a very few ML models have used more advanced neuroimaging parameters (11,23), and most have failed to incorporate patient-specific cerebrovascular features in their models, thus missing out on exploiting the rich vascular information (14,24,25) despite their significant impact on patient outcomes (2,9,26). Not surprisingly, the outcome prediction studies have reported a relatively low specificity and sensitivity with an area under the curve (AUC) for the receiver operating characteristic curve (ROC curve) under 0.76 (27,28).

We hypothesized that incorporating patient-specific cerebrovascular morphological features would improve stroke diagnosis and long-term outcome prediction. A major challenge in the automatic estimation of the CI and other features of cerebrovasculature is the lack of a validated method for vascular segmentation and feature extraction from baseline CTA or MRA scans. In the past few years, there have been significant advances in automatic cerebrovasculature segmentation methods, which refers to partitioning an image into multiple segments to separate the regions of interest from the background, i.e., “vessels” and “non-vessels,” by assigning a label to each image pixel, (29–31). We previously developed a novel, validated algorithm for accurate segmentation and geometric feature extraction of the cerebral vessel network (32). However, the segmentation must be extremely fast, and produce vessel network maps in real-time, for clinical applications. For this purpose, deep learning techniques are gaining popularity as they enable instantaneous 3D segmentation of volumetric imaging data. Most deep learning efforts in literature have used small datasets for neural network training and applied threshold-based vascular maps as the ground truth to generate binary vessel trees (30,33). These limitations lead to inaccuracies in the final segmentation due to insufficient training data, the inconsistent nature of the thresholding-based approach, and low inter-observer agreement in manually segmented ground truth.

In this work, we present a novel hybrid image processing and artificial intelligence pipeline for stroke patient triage. Using our validated segmentation algorithm as ground truth (32), we train a convolutional neural network (CNN) for instantaneous, accurate, and automatic segmentation of the vascular network from the raw CTA or MRA scans that aids in subsequent extraction of the complex geometric features of the cerebral vessel network, namely – total length, average diameter, branching pattern, total volume, vessel tortuosity, and fractal dimension. We then used these features in conjunction with our previously developed statistical cerebrovascular atlas (34) to automatically detect the presence and site of the vessel occlusion, and calculate the CI for each stroke patient. The findings were validated against ground truth clinical assessment and grading of each subject’s CI. Lastly, we developed a predictive ML algorithm to incorporate the automatically extracted cerebrovascular features and CI in combination with the commonly used clinical and imaging parameters to predict the 90-day functional outcome of AIS. With this work we aim to improve various aspects of stroke patient triage by bringing imaging data to the forefront of treatment decisions.

## METHODS

### Datasets

This retrospective study uses multiple anonymized datasets, each approved by the corresponding IRB. Table 1 lists the details of all the imaging data used in this study.

**Table 1.** The imaging datasets and the corresponding number of scans used in the various aspects of the study.

| Databases |  |  | Number of subjects |  |  |  |
| --- | --- | --- | --- | --- | --- | --- |
| Name | N (Female) | Resolution (mm<sup>3</sup>) | CNN<br>(train/ test) | Stroke<br>detection | CI<br>validation | Outcome<br>prediction |
| MIDAS (healthy) | 109 (57) | $0.5 \times 0.5 \times 0.5$ | 99/10 | 50 | - | - |
| OASIS3 (healthy) | 66 (29) | $0.3 \times 0.3 \times 0.3$ | 41/25 | - | - | - |
| Stroke | 100 (54) | $0.5 \times 0.5 \times 0.5$ | 0/10 | 100 | 56 | 100 |
| CTA (healthy) | 10 (4) | $0.43 \times 0.43 \times 0.62$ | 0/10 | - | - | - |

ToF-MRA was the modality used in all databases except the Healthy CTA database, where CT angiography was used. Additionally, 12 of the 100 stroke patients were excluded from the outcome prediction due to unavailability of their mRS outcome.

To train and test the CNN segmentation algorithm, we used ToF-MRA scans of 175 healthy subjects consisting of 109 subjects from the MIDAS public database (CASILab at the University of North Carolina, Chapel Hill, NC; distributed by Kitware, Inc.) and 66 subjects from the OASIS-3 study (35). We also obtained MRA scans of 100 AIS patients from the Centre Hospitalier Universitaire Vaudois in Lausanne, Switzerland. All patients had a confirmed diagnosis of AIS due to an occlusion in the anterior circulation in the internal carotid artery (ICA), the middle cerebral artery (MCA) at the M1, M2, or M3 segments, or a combination of these. The scans were performed within 72 hours from stroke onset. The database also included demographic, clinical, and additional neuroimaging data, including NIHSS, ASPECTS and perfusion mismatch volume and ratio, as well as 90-day functional status assessed using the modified Rankin scale (mRS), when available. We used the MRA scans and patient data to extract the patient-specific vascular features and collateral index and train our outcome prediction model. Out of the 100 AIS patients, 12 had to be excluded from the prediction model due to the unavailability of their mRS outcome.

### Segmentation and feature extraction

For CNN-based cerebrovascular segmentation, we adapted and optimized the U-Net architecture. The U-Net framework, presented by Ronnenberger et al. (36), has previously been successfully used for various medical image segmentation applications. Out of the 175 healthy MRA, 10 stroke MTA and 10 CTA scans, we used 140, 25, and 30 scans for training, validation, and blind testing, respectively with a stratified sampling of the CNN segmentation model. We used our validated methodology for segmentation and vascular feature extraction to obtain the “ground truth” of the segmented vascular maps and their respective features (32). The segmentation algorithm involves a multi-step process, resulting in a 3D binary volume of the vessel network, with being able to detect vessels with diameters as small as the imaging resolution. It was shown to perform better than other available segmentation algorithms under technical issues that can cause intensity inhomogeneities in imaging data. After segmentation, we performed a skeletonization of the vessel network and extracted the vascular geometric features, described in the *Supplementary Material*.

For CNN training, the raw scans were the model input and mapped to their corresponding segmented vascular networks, using the above method as output. The model architecture (Fig. 1, *Supplemental Materials*) includes skipping connections built in between the encoding and the mirrored decoding path for each scale level, with deconvolutional layers replacing the 2-stride convolutions. We use a small kernel size of 3×3, and our model consists of 18 total layers – 9 double convolution layers each in the encoding and decoding segments. We implement the Rectified Linear Unit (ReLU) activation function throughout the layers and a Sigmoid function at the last layer for the final prediction of grayscale pixel intensities between 0 and 1. Adam optimizer was used for the gradient descent, and the model was trained for 50 epochs until the loss minimized. Early-stopping and drop-outs were used to prevent overfitting and improve generalization (37). We defined an application-specific loss function known as “Dice Loss,” which, when minimized, maximizes the overlap of segmentation prediction and ground truth, measured using the Dice Similarity Coefficient “SoftDice” (38).

**Fig. 1.**
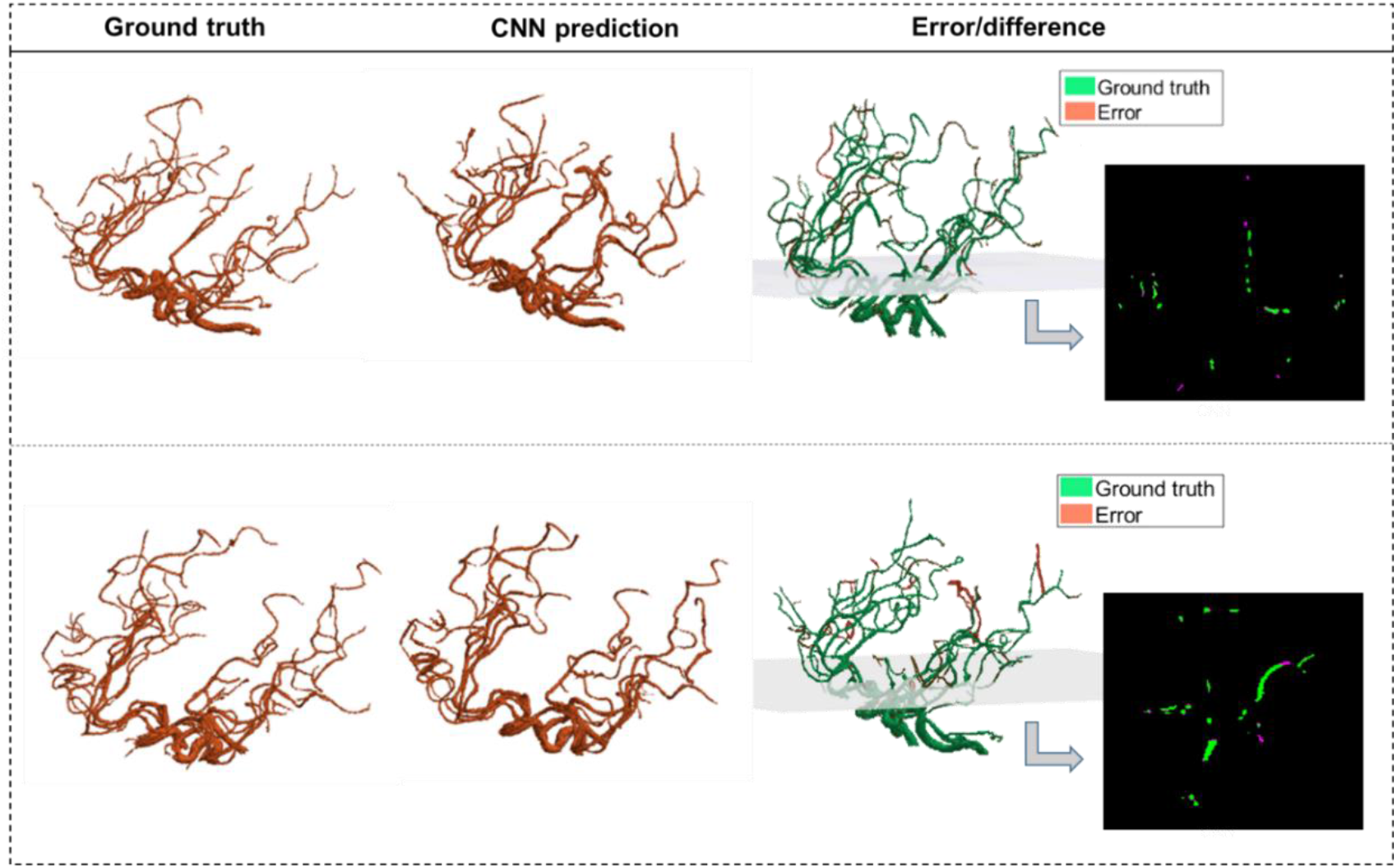
The CNN predictions of the segmented vascular maps. CNN segmentations of two MRA scans from test data are shown alongside the corresponding ground truth segmentations obtained using our validated algorithm. The third column shows the error volume overlaid on the ground truth volume, with one 2D panel showing the overlap between corresponding slices of the ground truth and the error.

For CNN validation and testing, we compared the model predictions against the vascular maps obtained by our segmentation algorithm (ground truth) using an image matching metric, namely the Dice Similarity Coefficient (DSC). The DSC quantifies the overlap between the ground truth and the prediction for each slice of the 3D volume as a measure of accuracy and is obtained by averaging the DSC per 2D cross-sectional slice across each 3D volume and then averaging across all the test scans. Also, as a further quantitative assessment of model performance, we compared the following extracted features obtained by the CNN model against the ground truth: total length, number of branches, total volume, and average diameter. Lastly, the CNN model was tested on the CTA scans of 10 healthy subjects and MRA scans of 10 AIS patients to accurately evaluate the model’s ability to segment vasculature in different modalities and health status.

### Stroke detection and occlusion localization

Of 100 AIS patients, the occlusion sites were the Internal Carotid Artery (ICA) in 18, tandem ICA-MCA in 17, and M1 and M2 segments of the MCA in 46 and 19, respectively. We utilized our previously developed probabilistic cerebrovascular atlas of spatially co-registered cerebral vessel maps of 175 healthy adults (34). The atlas was labeled to denote the five major vascular territories: (1) the ICA, (2 and 3) the left and right MCA, (4) Anterior Cerebral Artery (ACA), and (5) the posterior cerebral artery (PCA) and basilar artery (BA), as illustrated in Fig. 3.

We implemented a two-step approach for automatic stroke detection based on previous preliminary studies that showed a significant difference in cerebrovascular features between stroke patients and healthy subjects (32,34). In the first step, the presence of occlusion was determined by comparing the vessel density of the patient-specific vascular network with the cerebrovascular atlas using the total vessel length, volume, and the number of branches. A significantly lower vessel density, defined as more than three standard deviations below the average healthy subjects, indicated an occlusion. In the second step, each of the five vascular territories of AIS patients was compared with the corresponding territory in the atlas. The territory with the largest deviation score from the healthy average was identified as the occlusion location (Fig. 3). We also compared the vessel density between the left and right cerebral hemispheres for each patient for cases where the comparison against the atlas was not sensitive enough. The analysis was performed for all 100 AIS patients, and 50 randomly selected healthy controls to assess the algorithm’s ability to detect the occlusion’s presence and location. We evaluated the algorithm by calculating both steps’ sensitivity, specificity, and positive and negative predictive values (PPV and NPV).

### Automatic collateral index estimation

The collateral index (CI) quantifies the development of a patient’s collateral vessels. In clinical settings, CI is usually graded as either good, intermediate, or poor. The clinical grading which was utilized for developing and validating our method is as follows (39):

0 = collateral supply absent
1 = collateral supply filling >0% but ≤50%
2 = collateral supply filling >50% but <100%
3 = 100% collateral supply

Of the 100 AIS patients, we had access to the CI for 56, graded in the clinic by a neuroradiologist, since this metric is not routinely estimated in all stroke patients due to the time constraints and manual nature of the task. These clinically-evaluated CIs were used as the “ground truth” to develop and validate our algorithm. We then use the validated method, described below, to obtain the CI for the remaining 44 AIS patients automatically.

Using our cerebrovascular atlas (34), the relative vessel density in the collateral region was calculated for each patient compared to patients with the fully developed collateral network (CI=3). Based on this relative index obtained using linear regression, we grade the collaterals per patient on a scale of 0 (absent collateral supply) to 3 (100% developed collateral supply). We used this automatically estimated CI for the 100 patients as an outcome predictor in our ML model, given the well-established impact of collateral circulation on the patient’s response to ischemia and eventual treatment (15,16,40).

### Functional outcome prediction

The functional status of the AIS patients was evaluated at the 90-day mark using the modified Rankin scale (mRS) (Table 1, *Supplementary Material*) (41). The input data for the prognostication model comprised of patients’ clinical and imaging variables, including demographic information, pertinent past medical history, stroke symptoms and severity (the baseline NIHSS), and data from initial brain imaging studies, including the ASPECTS and perfusion metrics. A larger perfusion mismatch volume or ratio indicates a larger salvageable tissue that may be amenable to acute treatments (42–44).

Cerebrovascular geometric features have been shown to correlate with aging and pathologic states (32,34,45–47). To utilize the predictive ability of the rich imaging-based vascular information, we incorporated novel patient-specific cerebrovascular geometric features extracted by the segmentation algorithm described above (32) as well as automatically estimated CI into our ML model as additional predictors of the 90-day mRS. Table 2 in the *Supplementary Material* lists all the features used as predictors to train the model.

**Table 2.** CNN model performance on test data per geometric feature between the predictions vs. ground truth

| Validation metric | Value (mean ± std) |
| --- | --- |
| Avg. DSC | $0.94 \pm 0.05$ |
| Avg. error in total length (%) | $3.40 \pm 0.31$ |
| Avg. error in the number of branches (%) | $1.90 \pm 0.44$ |
| Avg. error in total volume (%) | $3.18 \pm 0.26$ |
| Avg. error in average diameter (%) | $2.11 \pm 0.10$ |

The in-clinic assessment of the mRS at 90 days post-stroke was considered the ground truth, trichotomized into good (mRS 0-1), moderate (mRS 2-3), and poor (mRS 4-6) outcome groups. Reducing the 7-class mRS to the three output classes facilitates prediction and provides a more granular classification than the dichotomized predictions in literature (14,20,25). We used supervised ML to train the predictor model using the Classification Learner app in the MATLAB® package (Mathworks, MA). The dataset was divided into 80% training and 20% test datasets for validation. We also included stratified sampling and five-fold cross-validation to improve learning and prevent overfitting. The features were then ranked using the chi-square test based on the univariate associations between each categorical or continuous predictor variable and the 90-day mRS outcome. The ranked features were used as predictors, and the three final outcome groups were the ground truth for the output classes. The final outcome prediction model was chosen based on the training and validation performance metrics. The model performance was assessed by calculating the accuracy using the true positive and false negative rates and the positive and negative predictive values. The findings were visualized by the AUC of the ROC curve. Since we are performing a multi-class prediction, the AUC was computed for the ROC curves using the one-against-rest method for multi-class models (49).

## RESULTS

### CNN segmentation model

The average Dice Similarity Coefficient (DSC) between the ground truth segmentation and CNN predictions across the multi-resolution test data was 0.94. The model captured the vascular branches and preserved the vascular volume, with the average error margins under 4% for all geometric features: total length (3.4%), number of branches (1.90%), total volume (3.18%), and average diameter (2.11%). Fig. 1 shows two predicted segmentation maps and their corresponding ground truth. The quantitative measures of model performance on the test dataset are shown in Table 2.

### Stroke detection and occlusion localization

Our model identified 92 strokes among 100 AIS patients and 50 healthy subjects with a sensitivity of 92% and specificity of 94%. The algorithm only missed the presence of an LVO in 8 cases due to the overall vessel length and volume not being significantly different than the atlas due to the presence of significant vessel density in the proximal part of the middle cerebral artery (MCA) in case of M2 strokes, in the distal segment of the MCA (Fig. 2). Additionally, the algorithm detected three false positives among the 50 healthy subjects due to the vascular networks deviating significantly from the atlas.

**Fig. 2.**
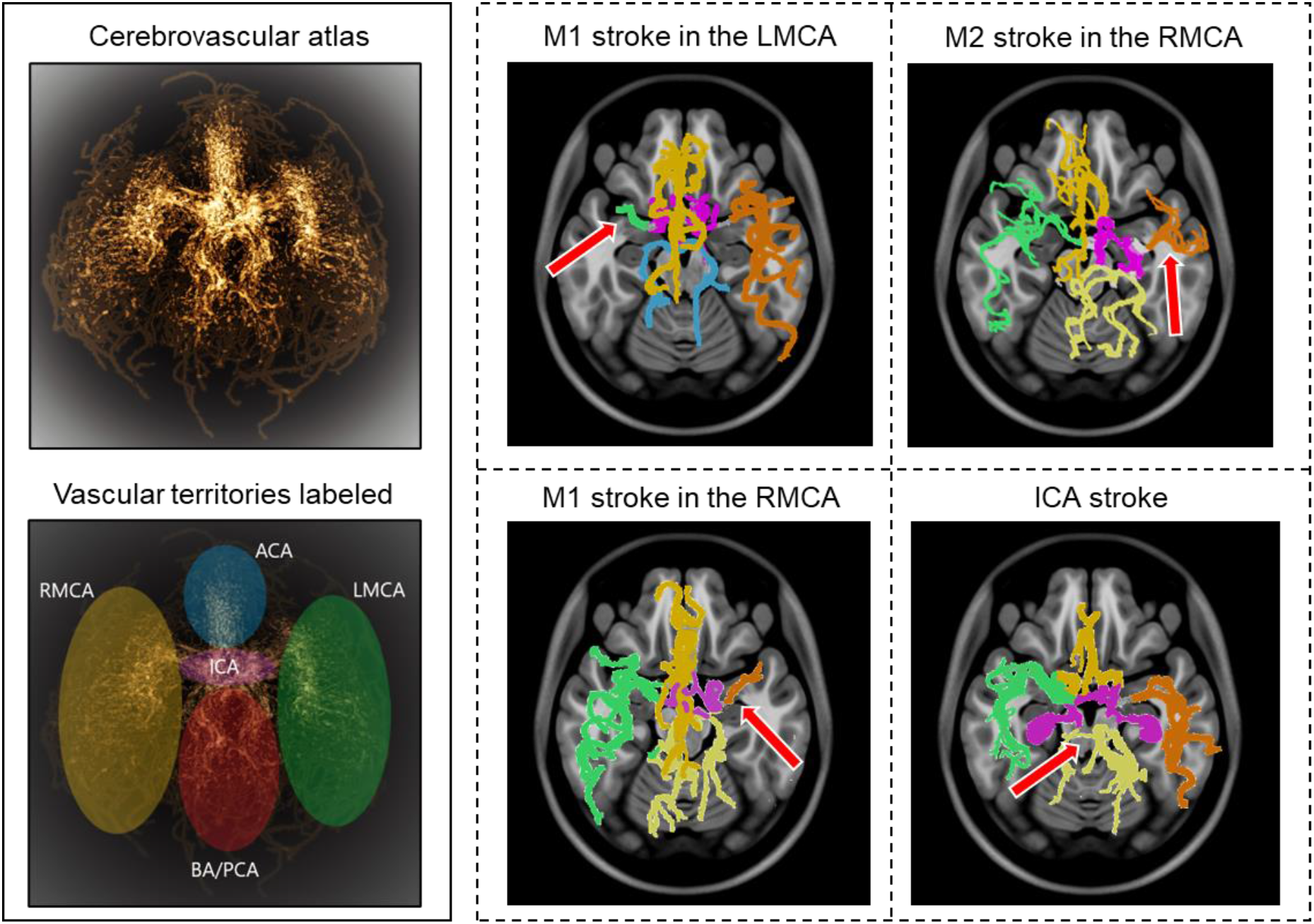
Stroke detection and occlusion localization. The brighter regions of the 3-dimensional probabilistic atlas (top-left) show maximum intensity projection and correspond to a higher probability of vessel occurrence. The brain vascular was divided into five major territories, illustrated in different colors (bottom-left). The Cerebrovasculature map of four stroke patients is shown on the right. The red arrow indicates the location of the occluded vessel

The model had an accuracy of 95.6% in identifying the region containing the occlusion, with only four mis-localizations out of the 92 stroke cases identified. These errors are due to outliers in vascular patterns deviating from the average atlas. Overall, the model achieved a 95.56% positive predictive value (PPV) and a 95.56 negative predictive value (NPV).

### Collateral index (CI) calculation

The algorithm correctly estimated the CI in 49 out of the 56 strokes MRA, yielding a sensitivity of 87.2%. Fig. 3 shows the vascular tree, extracted vessel network from the ipsilateral collateral region, and the estimated CI for 4 cases, each corresponding to a CI score from 0 to 3.

**Fig. 3.**
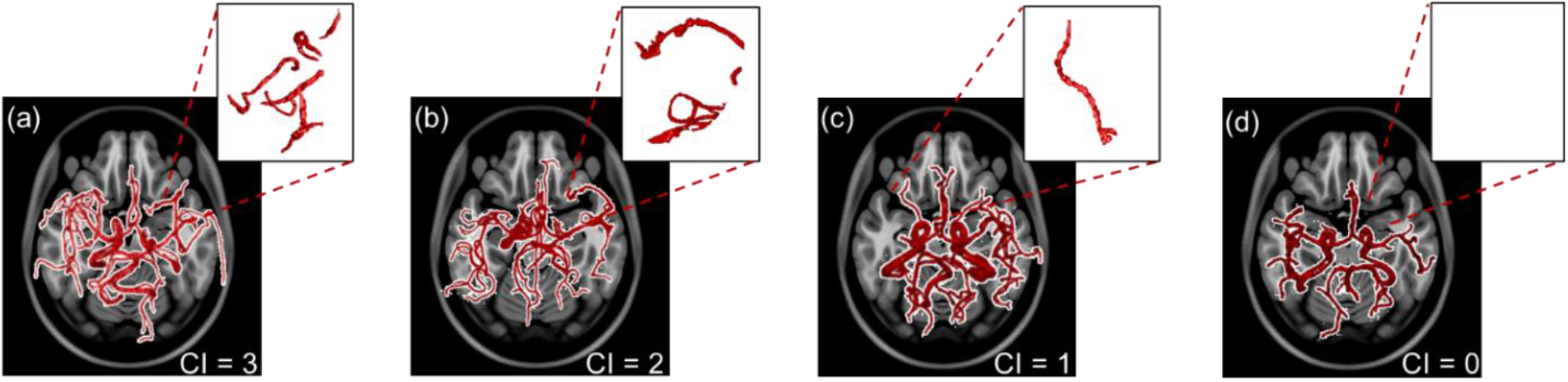
The extracted vascular networks and estimated collateral index for the four varying levels of development of the collateral network in patients with a middle cerebral artery stroke. The collateral index (CI) is shown at the bottom right of panel a-d. The top right sub-panels show the vessels in the corresponding collateral region of the ipsilateral hemisphere.

### Functional outcome prediction

Fine Decision Trees were adopted for multi-class prediction of the 90-day modified Rankin scale (mRS) (41). The basic predictor model using conventional predictors of functional outcomes, reached an area under the curve (AUC) of the receiver operating characteristic (ROC) curve of 0.63±0.01 (Fig. 4-a), similar to currently available models. By including the automatically estimated CI in the prediction model, the and the AUC of the ROC curve increased to 0.74±0.02. The additional incorporation of vascular geometric features further increased the prediction accuracy with an AUC of the ROC curve of 0.83±0.02 (Fig. 4-a). ROC curves in Fig. 5 are overlaid on the same graph to highlight the AUC values, with the true class being the ‘Good’ outcome (i.e., mRS 0-2). The confusion matrices (Fig. 4-b and 4-c) show the prediction accuracy per outcome group for the predictive models with and without the morphologic features.

**Fig. 4.**
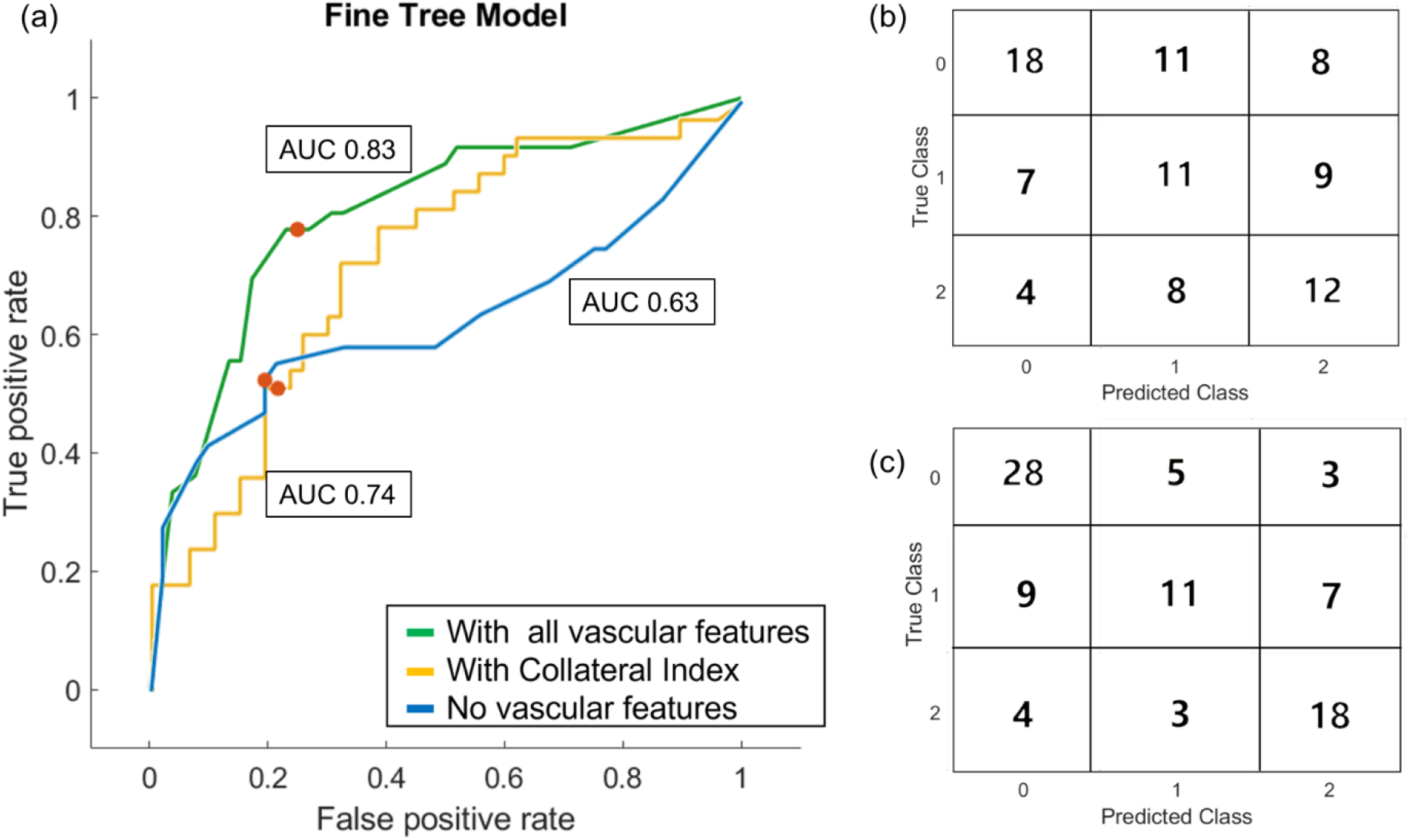
Results of the outcome prediction models. (a) ROC curves for the three Fine Decision Tree models are plotted. All three models included the baseline clinical and imaging data. AUC-ROC for the model without any vascular features was 0.63. AUC-ROC increased to 0.74 in the model trained with the collateral index but without other geometric features. Incorporating all vascular features improved the model’s performance with AUC-ROC =0.83. The confusion matrix for the prediction model before (b) and after (c) incorporating the vascular geometric features and auto-estimated collateral index are shown.

## DISCUSSION

ML strategies have been used in various applications in stroke medicine (9,14). In this work, we presented an end-to-end automatic ML approach for stroke triage, consisting of a CNN-based cerebrovascular segmentation and morphologic extraction, an automated algorithm for stroke detection and collateral circulation assessment, and finally, a 90-day functional outcome predictor.

### Cerebrovascular segmentation using CNN

Accurate and efficient segmentation of brain vascular imaging by extraction and visualization of the 3D cerebrovascular network is critical for clinical practice. We had previously developed and validated a method to detect brain vessels as small as the image resolution (voxel size) with superior performance compared to other freely available segmentation software (32). However, depending on the image resolution and computing resources, our method took up to 20-30 minutes to segment one vascular imaging study. Here, we used the segmented vascular maps extracted by our algorithm as the ground truth to train a U-Net architecture-based CNN model for accurate and instantaneous segmentation of 3D vessel networks from raw imaging data. The existing CNN-based segmentation methods require manual annotation with increased “noise” in the final segmented map pertaining to the erroneous prediction of non-vessel voxels from the skull or bright spots on the scans (30,33). Our model demonstrated accurate segmentation without requiring manual annotations. It detects the finer vessels present in the ground truth segmentation with a small error margin and, in some cases, detects smaller tapering vessel voxels that were not present in the ground truth segmentation of the blind test data due to poor contrast in the raw imaging scan at those voxels. The segmented vascular edges are smooth, and the CNN predictions do not miss vessel pixels around the boundary surface of the vessel cross-sections, as seen in 2D slices, resulting in the diameter and other geometric features to be computed accurately.

### Stroke detection and occlusion localization

Vascular morphology differs significantly between healthy and stroke subjects (32,34). Implementing complex cerebrovascular features and quantitative measurement of deviation from the average healthy atlas forms the basis of our stroke detection algorithm. Using a labeled atlas of healthy vessel networks and their inherent geometrical properties, we identified the anatomical region of occlusion in the most commonly occurring ischemic strokes. The sensitivity of our algorithm is comparable to (and in some cases higher than) previously published methods, and the specificity has improved as well (4,5). The high sensitivity and specificity of the stroke detection algorithm demonstrate the applicability of vascular geometry in automated stroke diagnosis and occlusion localization rather than a simplistic hemispheric comparison that may lead to false detections due to inconsistent vascular symmetry between the two hemispheres. Early automatic diagnosis of AIS and identifying the occluded could be invaluable in radiological screenings in case of an emergency or lack of on-call neuroradiologists in smaller medical centers.

### CI estimation and functional outcome prediction

A large number of studies support the significant benefits of EVT in treating acute ischemic stroke (1,2,10,11,50). The eligibility for EVT is expected to expand, with a shift from rigid time-based treatment protocols to imaging-based strategies that incorporate patient-specific factors into therapeutic decision-making, such as collateral circulation (2). Variations in vascular anatomy affect cerebral hemodynamics and even rates of neuronal degradation (51)(52) during ischemia and, thereby, response to treatment. A more developed collateral circulation provides more time for therapeutic interventions and impacts clinical outcomes (15,16). Therefore, a better understanding of each patient’s cerebrovasculature and collaterals are pivotal to expanding eligibility for acute treatments (17). The method presented in this study can accurately, rapidly, and automatically calculate the CI. This development can drastically impact patient triage and reduce the time for diagnosis and treatment (53).

Prognostication of AIS remains challenging (24) despite its tremendous impact on decision-making for patients, their families, clinicians, and society (27,54). An algorithm that can achieve early, reliable, and accurate prognostication is lacking. Many previous attempts at using ML for outcome prediction after AIS have yielded low-performance algorithms with low sensitivity and AUC for ROC curves under 0.76 (24,25,28,54).

Vessel structure and geometry, including lumen diameter and branching patterns, are known to impact the patient’s response to ischemia and reperfusion (12,15,48). This study presents a novel ML approach incorporating quantitative cerebrovascular information to predict the functional outcome. Adding the automatically graded CI as a predictor significantly improved the 90-outcome prediction, shown as a 17% higher AUC of the ROC curve. Including other vascular geometric features further enhanced the predictive utility of the algorithm, as shown in Fig. 5.

### Limitations

The stroke detection algorithm calls for a comparison with the cerebrovascular atlas, which, in turn, requires the patient-specific vascular network to be spatially co-registered to the Montreal Neurologic Institute (MNI) space, utilized to normalize all patient scans spatially. However, the standardized MNI atlas space is intended for MR scans, and CT scan images cannot be co-registered to the same space. Thus, for a wider appositeness of this method, CT scan data needs to be registered to this common space. Additionally, due to the inherently distinct nature of subject-specific vascular anatomy, patient vascular network alignments sometimes differ significantly from the probabilistic atlas in the cartesian space. This can cause errors in stroke detection as well as collateral estimation algorithms.

As a pilot study, we retrospectively analyzed a smaller stroke patient database (n = 100) to establish our methods and the utility of cerebrovascular morphology in stroke diagnosis and prognostication. Outcome prediction typically requires a large and representative patient database to assess predictive features accurately. Our training dataset was also limited to patients with large vessel occlusion in the anterior circulation. Furthermore, patients with missing outcomes were excluded from the final model (n = 12). A future large and prospectively collected dataset of patients with various stroke syndromes, including those with strokes in the posterior circulation and medium or small vessel occlusion, is necessary to solidify the effectiveness of vascular geometry as a predictive tool in AIS patients.

## Conclusion

We presented a novel end-to-end quantitative machine-learning strategy to extract patient-specific cerebrovascular morphology accurately, rapidly, and automatically from segmented vessel trees, automatically detect and localize LVOs, calculate the collateral circulation index, and predict 90-day functional outcomes. This approach aims to improve the accuracy and efficiency of detecting and localizing LVO and the fidelity of predicting the functional outcomes of stroke. Our method for automatic CI grading can help address the incongruity between the significant impact of collateral circulation assessment in AIS patients and the lack of time and resources to perform this task in the acute hospital setting. Through this approach, we highlight the need for patient selection and treatment decisions to be based on quantitative, imaging-based information along with clinical patient evaluation.

## Data Availability

The code and materials used in the analysis can be made available upon request. Two of the databases used in the study are publicly available.

