## Supplementary material for "End to end stroke triage using cerebrovascular morphology and machine learning"

**Detailed Methods**

Datasets:

An Allegra 3T MR scanner (Siemens Medical Systems Inc., Germany) was used for data acquisition in MIDAS database with 0.5 mm^3^ resolution. The scans in the OASIS-3 database were acquired on three different Siemens scanner models: a 1.5T scanner with a 16-channel head coil and two 3T scanners with a 20-channel head coil with 0.3 mm^3^ resolution. Furthermore, Stanford University School of Medicine provided head CTA scans of 10 healthy subjects, acquired on a GE Lightspeed scanner at 100-120KV after a bolus injection of 90–120 ml contrast media (Isovue-370 mg/ml) at the injection rate of 4–5 ml/s with 0.5 mm^3^ resolution. The stroke MRA scans acquired on a 1.5T Siemens scanner with 0.5 mm^3^ resolution

Multiple datasets were used in the various parts of this study – 175 MRA scans of healthy subjects, 10 MRA scans of stroke patients and 10 CTA scans of healthy subjects to train and test the Convolutional Neural Network (CNN) model; 100 MRA scans of stroke patients and 50 MRA scans of healthy subjects to develop and validate the stroke detection algorithm; 56 MRA scans of stroke patients to develop and validate the CI estimation algorithm; and lastly, extracted data and from 100 MRA scans of stroke patients for the outcome prediction model development and testing.

For the CNN model development, we used 140 MRA scans of healthy subjects for training and tested the model on four separate datasets consisting of 55 subjects: 35 MRA scans of healthy subjects (10 from the MIDAS dataset with 0.5 mm^3^ resolution and 25 from the OASIS3 dataset with 0.3 mm^3^ resolution), 10 MRA scans of AIS patients with 0.5 mm^3^ resolution, and 10 CTA scans of healthy subjects with 0.5 mm^3^ resolution (Table 1).

Vascular feature extraction:

For skeletonization of the segmented vessel network and subsequent feature extraction, a ‘branching node’ of the vascular tree, defined as a point connected to 3 other points (*i.e.*, a bifurcation), was used to calculate the global geometric features as follows:

1. Total length: The total length of the vessel network is calculated by summing the skeletal segments' length between 2 nodes.

2. Total number of branches: A ‘branch’ was defined as a sequence of points along the vessel beginning at a bifurcation point and ending either at the next bifurcation or the last point on the vessel (*i.e.*, a terminating branch).

3. Average and maximum branch length: All network branches' mean length (geodesic distance) and the longest branch found.

4. Average diameter: The mean diameter at all points on the centerline.

5. Total volume: Volume of the vessel network is calculated by considering the vessels as cylinders with varying diameters along the total length.

6. Fractal dimension: The fractality of the vessels was determined using the box-counting method. This feature is a measure of morphological complexity in cerebral vasculature.

7. Vessel tortuosity: Vessel tortuosity was defined using the sum of angles measurement between sets of 3 points on the centerline divided by the total length.

**Supplementary Table 1.** **The Modified Rankin Scale** used in clinics to assess functional outcomes in stroke patients at 90 days post recanalization. This is the final output from the prediction model to quantify patient status.

| **MODIFIED RANKIN SCALE** | |
| --- | --- |
| **0** | No symptoms |
| **1** | No significant disability despite some symptoms, able to perform all activities |
| **2** | Slight disability, unable to perform all activities but can function without assistance |
| **3** | Moderate disability requires some help, can walk without assistance |
| **4** | Moderately severe disability, unable to walk without assistance, unable to attend to bodily needs without assistance |
| **5** | Severe disability, bedridden, incontinent, requires constant nursing care and attention |
| **6** | Death |

**Supplementary Table 2**. **Clinical and imaging variables used to train the prediction models**

| **Clinical and demographic information** | **Imaging-based clinical features** | **Vascular geometric features** |
| --- | --- | --- |
| Age | ASPECTS score | Total length |
| Sex | Side of the stroke | Number of branches |
| Baseline NIHSS | Occlusion location | Total volume |
| History of stroke | Perfusion mismatch ratio and volume | Average diameter |
| Smoker (y/n) | Collateral Index (CI) – ground truth | Tortuosity |
| Diabetes mellitus (y/n) |  | Fractal dimension |
| Hypertension (y/n) |  | Auto-estimated CI |

The cells in blue represent the conventionally used features, yellow refers to the automatically estimated CI using our method and green cells represent the novel vascular predictors.

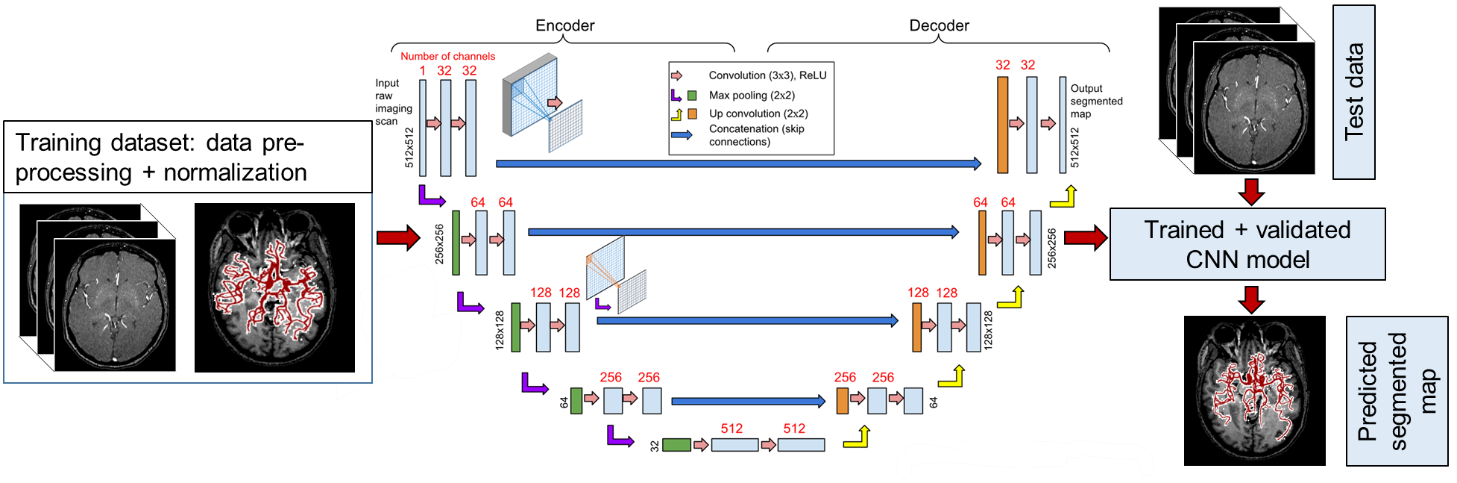

**Supplementary Fig. 1.** **CNN-based segmentation model.** The model is designed using the U-Net architecture, consisting of 18 total double convolution layers, 9 in each encoding and decoding segment, to achieve instantaneous segmentation.
